# Predictors of advanced-stage presentation among breast and cervical cancer patients in Ethiopia

**DOI:** 10.1101/2023.12.16.23300078

**Authors:** Birtukan Shewarega, Sefonias Getachew, Nigussie Assefa, Abdu Adem Yesufe, Josephin Trabizsch, Yonas Dandena, Biruck Gashawubeza, Adamu Addissie, Eva Johanna Kantelhardt, Muluken Gizaw

**Author notes:** Corresponding author information: Name: Muluken Gizaw, Address: Addis Ababa, Ethiopia, Tele.

## Abstract

Breast and cervical cancers are the most common causes of cancer incidence and mortality in women in Africa. Women with breast and cervical cancers in Sub-Saharan Africa (SSA) are frequently diagnosed at advanced stages. Delays in health seeking, diagnosis and treatment are contributing factors to high mortality in Ethiopia. This study aimed to assess predictors of advanced stage presentation among breast and cervical cancer patients attending public hospitals in Addis Ababa, Ethiopia. A cross-sectional study was conducted with a total of 418 patients at Tikur Anbessa specialized hospital and Saint Pauls’ Hospital Millennium Medical College from October to November 2021. Stages III and IV were considered advanced stages. Data were collected by reviewing medical records and in face-to-face interviews with a structured questionnaire. Bivariate and multivariate analyses were performed to examine the association between independent and outcome variables. A total of 269 patients with breast cancer and 149 patients with cervical cancer were included in the study, and the mean age was 44 years (SD = 10.9 years) and 50 years (SD = 11.2) years, respectively. About 66.9% of breast cancers and 71.1% of cervical cancers were diagnosed at an advanced disease stage. Rural residence (AOR = 2.041, 95% CI: 1.108–3.758), indirect referral (AOR = 3.8, 95% CI: 1.485–9.946), financial difficulty (AOR = 10, 95% CI: 1.859–56.495) and no prior advise/awareness about screening (AOR = 4.029 95%CI: 1.658–10.102) were independent predictors of advanced-stage presentation. This study revealed a high prevalence of advanced-stage breast and cervical cancer diagnosis in Ethiopia, similar to data collected 10 years ago, despite the introduction of a cancer control plan in 2015. For better implementation, interventions should aim to improve referral pathways, adapt screening and early detection services and increase cancer awareness at the community level in a culturally accepted way.

## Background

Breast and cervical cancers are the most commonly diagnosed cancers in women worldwide [1]. Women from Sub-Saharan Africa are disproportionally affected by both cancers, which contribute to the high burden of mortality in this region [2]. Both cancers are responsible for 50% of the cancer burden in women in Ethiopia [2]. Women with breast and cervical cancers in Sub-Saharan Africa (SSA) are frequently diagnosed at an advanced stage [3–5].

The five-year survival of cervical cancer in SSA is less than 50%, and an advanced stage at diagnosis and late initiation of treatment are the two outstanding contributing factors to poor survival in this region [6]. Breast cancer patients also present at an advanced stage due to the longer patient interval and lack of access to health facilities with diagnostic capacity in the patient’s vicinity [7]. Moreover, complicated referrals and longer diagnostic intervals have contributed to the delay in diagnosis and initiation of curative treatment [4].

Delays in referring these patients to the higher level are one of the contributing factors to late diagnosis and poor survival in SSA, including Ethiopia [8]. Lack of health infrastructure, a large patient flow in the hospital, long diagnostic intervals and patients’ repeated visits to different health facilities for diagnosis and treatment have been identified as contributing factors to late-stage diagnosis and poor survival in SSA [9–13].

However, despite a robust search of previous studies, we could not determine how delayed referral of those patients to secondary care is associated with advanced stage among both breast and cervical cancer patients in two public oncology centres in Addis Ababa, Ethiopia. Therefore, the present study explains the association between the direct referral path and advanced-stage presentation. We also sought to identify common individuals, health systems and clinical factors contributing to late-stage diagnosis among breast and cervical cancer patients diagnosed at a late stage.

## Methods

### Study design and Setting

An institutional based, cross-sectional study was conducted in Tikur Anbessa specialized hospital (TASH) and Saint Pauls’ Hospital Millennium Medical College (SPHMMC), two public referral hospitals (TASH and SPHMMC) in Addis Ababa. Tikur Anbessa Specialized Hospital is the largest cancer treatment centre including radiation treatment. St. Paul’s Hospital Millennium Medical College is the second referral and teaching hospital offering cancer treatment following TASH.

### Study participants

A total of 418 histopathologically confirmed breast and cervical cancer (aged >18 years) patients who were diagnosed at least 1 month prior to the data collection period and on follow up for treatment were enrolled in the study. Those breast and cervical cancer patients who were critically ill or missing dates regarding pathological diagnosis, cancer staging and symptom onset were excluded from the study.

### Sampling technique and sample size

The hospitals were selected purposively by the criteria of having oncology services. Consecutive sampling was used to enrol study subjects. The sample size was calculated using the single population proportion formula with Epi info version 4.1 software using the parameters 95% confidence interval, 80% power, 5% margin of error and proportion advanced-stage presentation of cervical cancer patient study conducted at TASH p = 55.2% [14].

### Variables and measurements

The following Aarhus statement criteria were used for classifying patient and diagnostic intervals:

The **patient interval/appraisal interval** was defined as the time interval from first symptom recognition (the time point when first bodily changes and/or symptoms were noticed) to presentation to the first health care provider, and a **patient appraisal /patient delay** was defined as a patient interval greater than three months (> 90-day)[15].

The **diagnostic interval** was defined as the interval from the date of first clinical presentation to the date of pathological diagnosis. The date of diagnosis was taken from the patient’s pathology report. A **diagnostic delay** was defined as a time interval between the first clinical presentation and the date of pathological diagnosis (diagnostic interval) greater than 30 days [4].

**Direct referral** occurred when patients with a given set of symptoms were referred within two weeks and obtained a diagnosis of cancer within two weeks in the first receiving facility [16].

**Financial difficulty** was defined as financial hardship incurred for transportation and medical expenses.

### Data collection and data quality assurance

Data were collected by a review of medical records and face-to-face interviews using structured questionnaires. Nurses who were working in the oncology department collected data, and training was offered for three days. Pretesting was performed prior to data collection in 20 respondents.

### Data analysis

Descriptive statistics were used to measure the frequency and proportion of advanced disease presentation. Bivariate and multivariable logistic regression analyses were used to identify predictors of advanced stage diagnosis.

### Ethical consideration

Ethical clearance was secured from the Institutional Review Board of the School of Public Health, College of Health Science, and Addis Ababa University (SPH/1119/2021). Verbal consent was obtained from all study participants. The confidentiality of participants was kept by not sharing their personal identifiers in all research processes.

## Result

### Socio-demographic characteristics of the respondents

The mean age was 44 years (SD = 10.9 years) and 50 years (SD = 11.2) for breast cancer and cervical cancer, respectively. Around 234 (56%) of the study participants were from urban areas, while 44% were rural dwellers. A quarter (106; 25.4%) of participants could not read or write, and 185 (44.2%) had attended secondary school or above. Around 267 (63.9%) respondents were married and 310 (74.2%) were Orthodox Christian by religion.

**Table 1.**
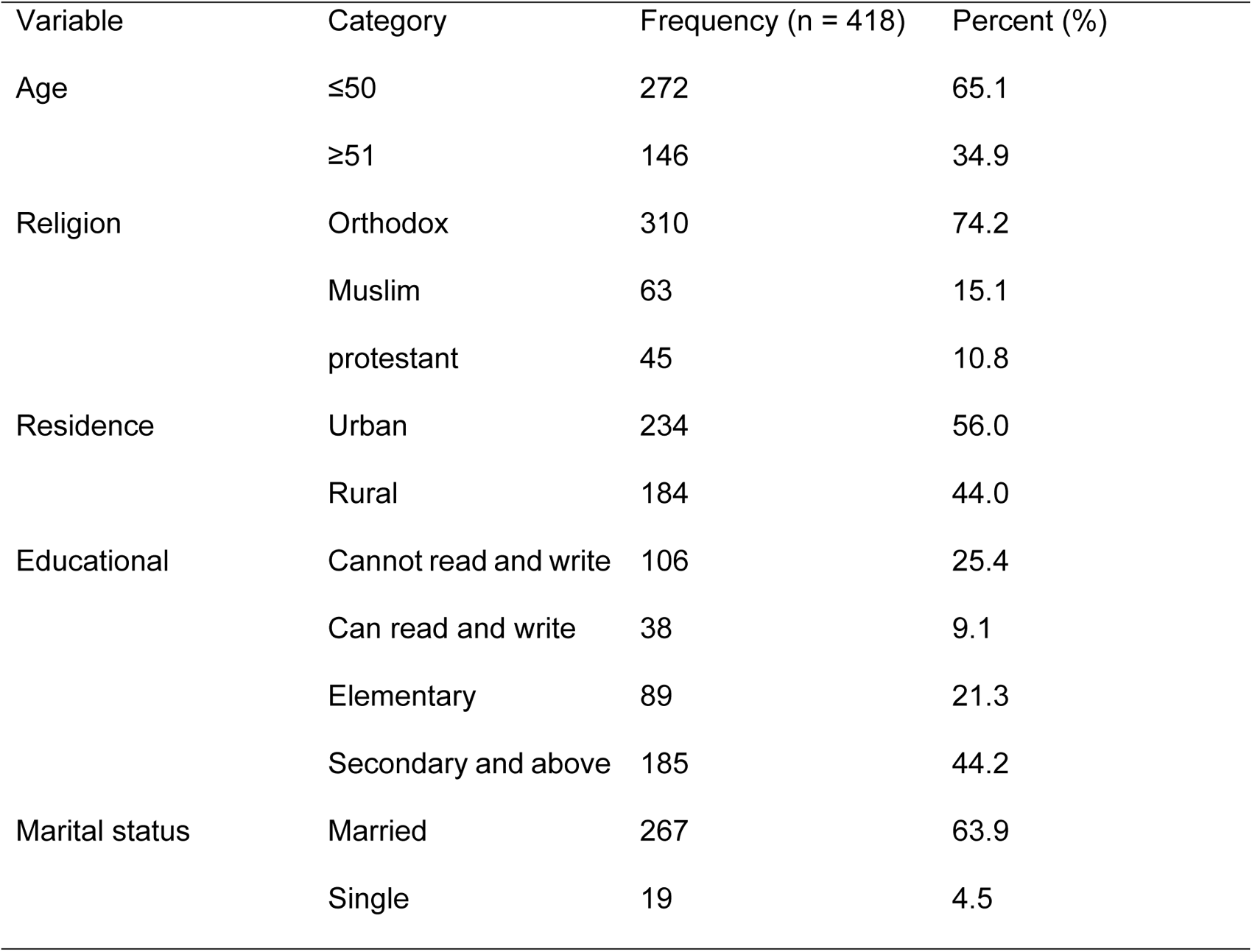
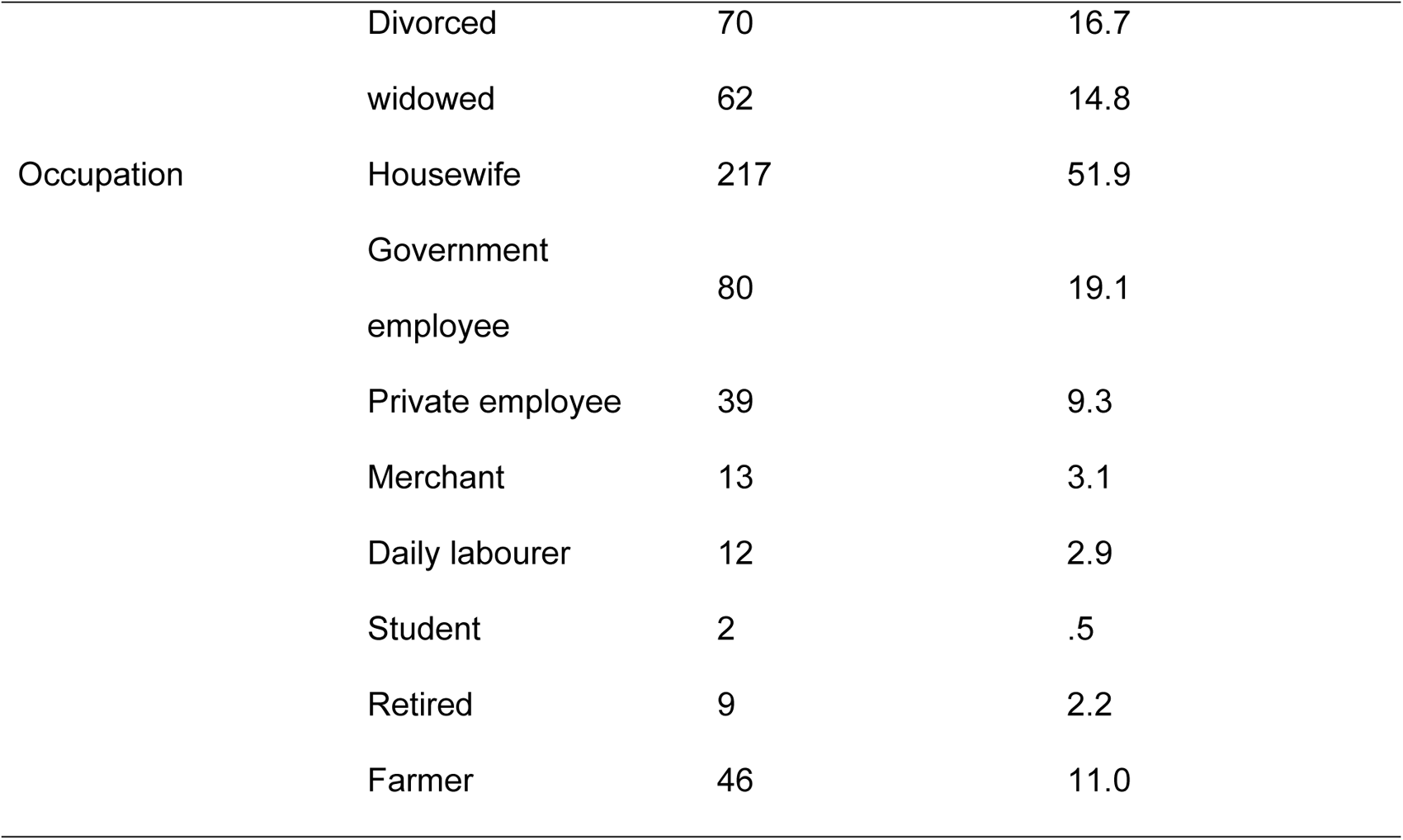
Socio-demographic characteristics of respondents in TASH and SPHMMC, 2021.

Close to half (132; 49.07%) of breast cancer patients ≤50 years of age had advanced-stage presentation, whereas only 48 (17.84%) respondents in the >50 age group had advanced-stage presentation. However, there was an equal proportion of cervical cancer patients with advanced-stage presentation, 53 (35.57%), in both age groups.

Around 36.43% of breast cancer and 34.23% of cervical cancer patients had a monthly income of less than 5000 ETB.

**Table 2.**
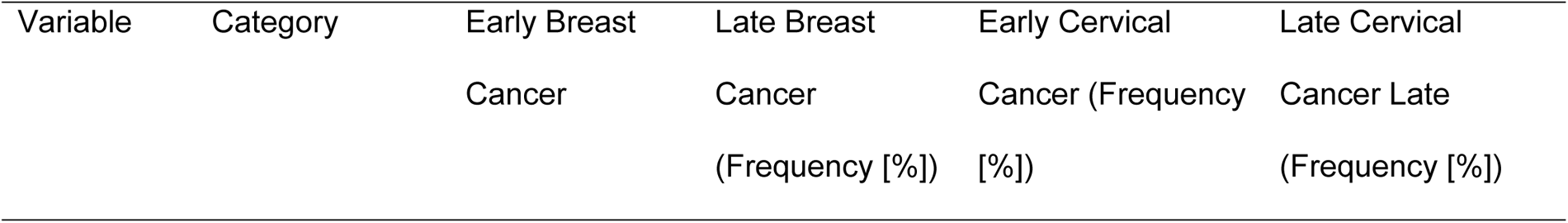
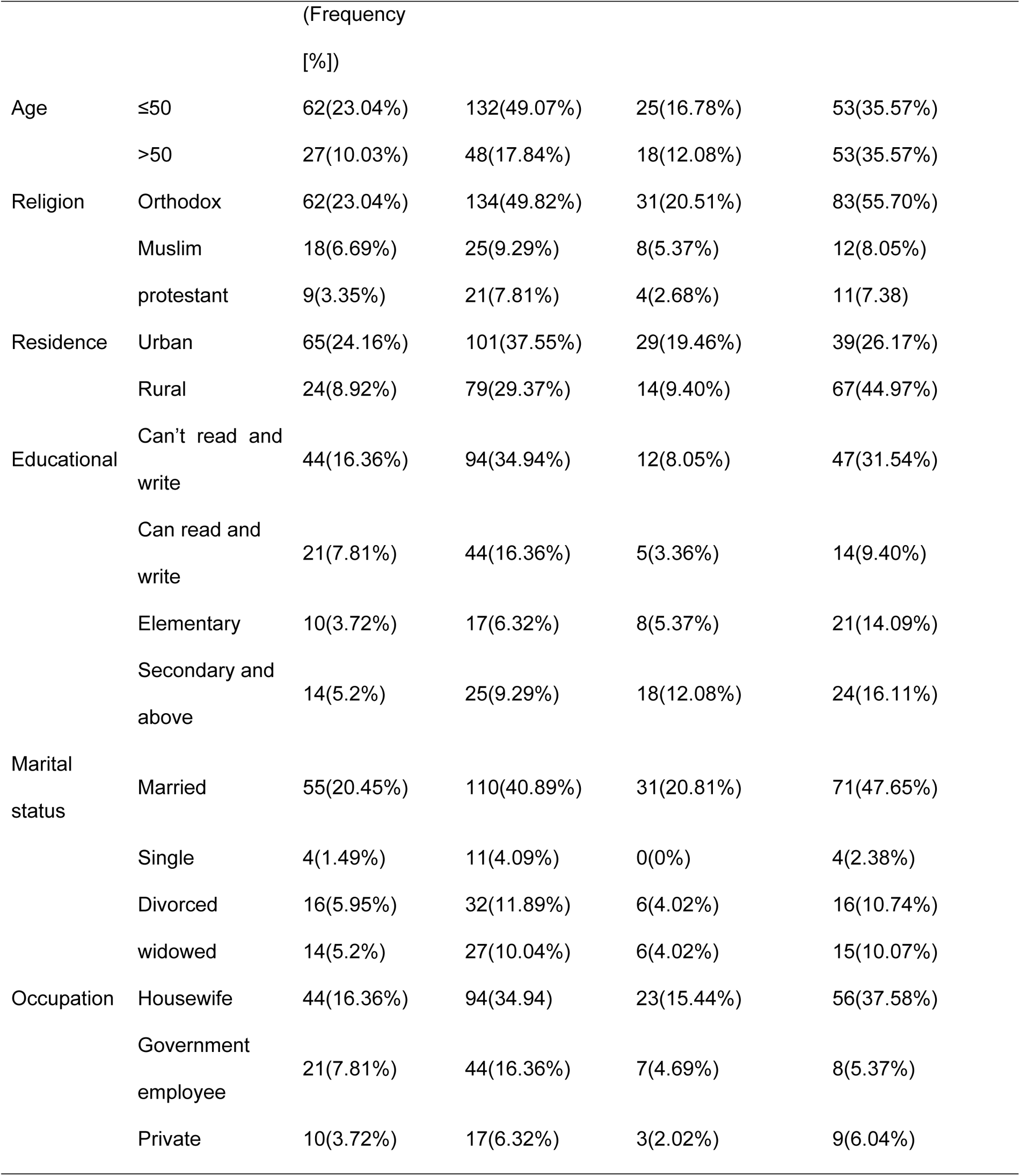
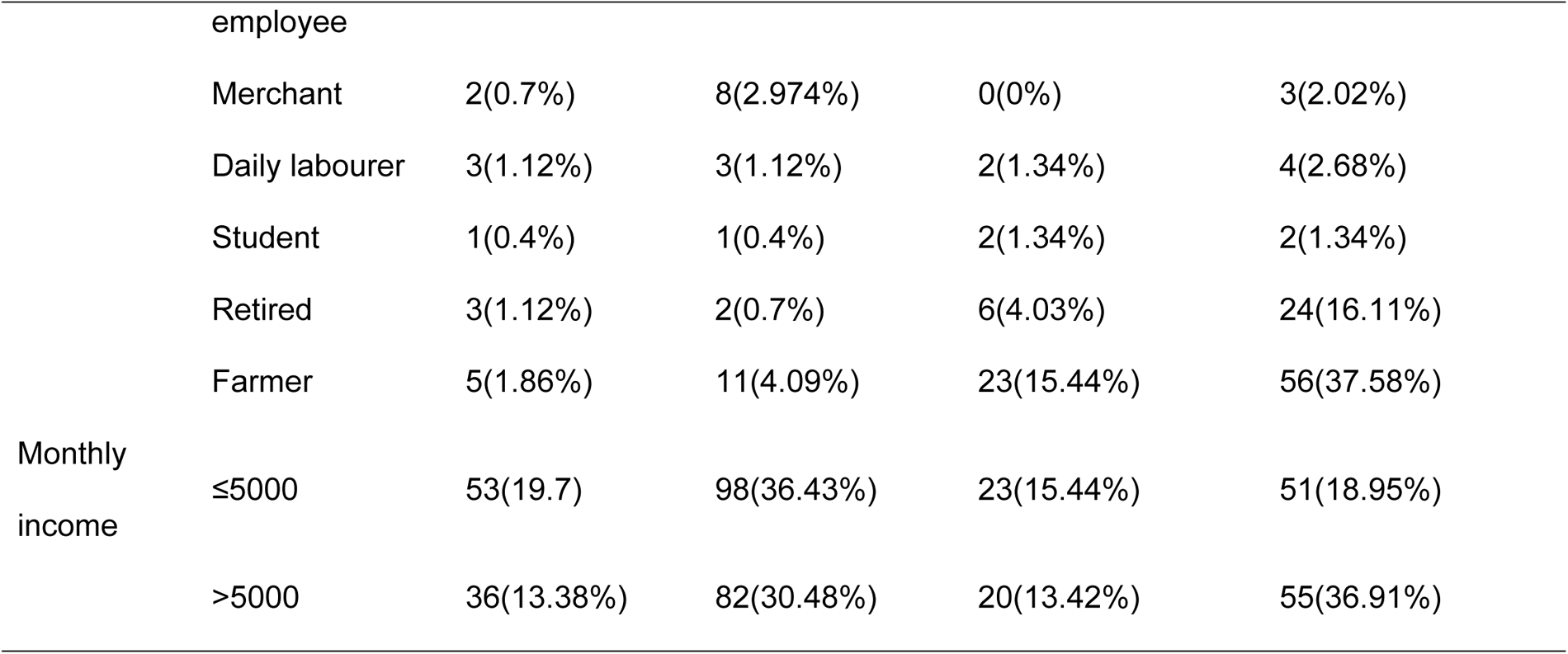
Socio-demographic characteristics of participants according to disease presentation at diagnosis in TASH and SPHMMC, 2021.

### Clinical characteristics of respondents

About half (51.3%) and 71 (47.7%) breast cancer and cervical cancer patients were on chemotherapy alone, respectively. About 38.3% of cervical cancer patients took chemotherapy and radiation therapy. The majority of breast cancer patients (66.9%) were diagnosed at an advanced disease stage (III & IV). Similarly, 71.1% of cervical cancer patients were in an advanced stage. Screening awareness was present in 108 (40.1 %) and 43 (29.8%) breast and cervical cancer patients, respectively. Among them, only 0.4% of breast and 6% of cervical cancer patients had a history of screening practice before diagnosis. Almost 85% of breast cancer patients had complained of a breast lump at presentation. Similarly, 92 (62.2%) cervical cancer patients had a presenting complaint of bloody vaginal discharge followed by pelvic pain (22.3%) or non-bloody vaginal discharge (10.8%).

**Table 3.**
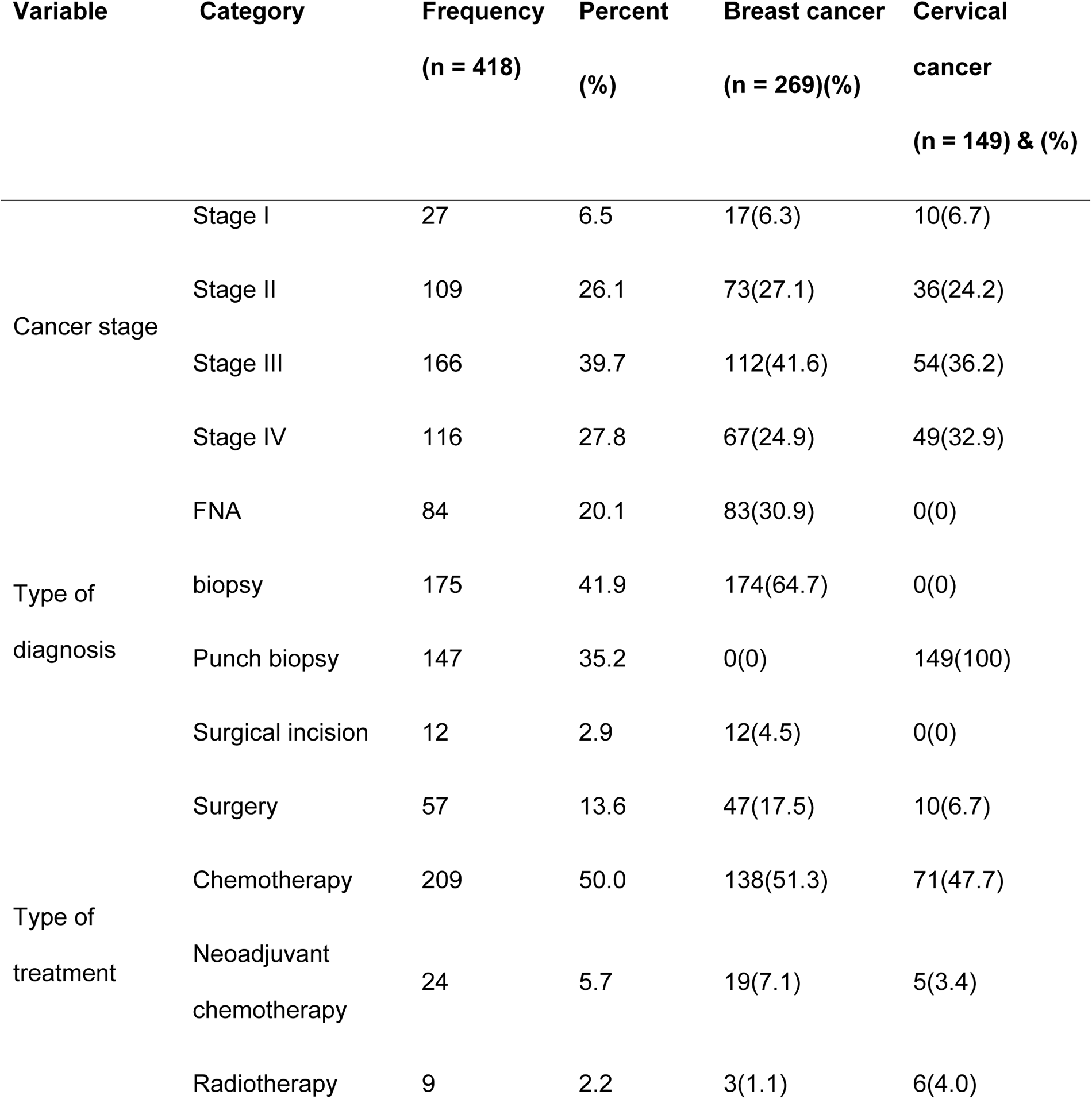
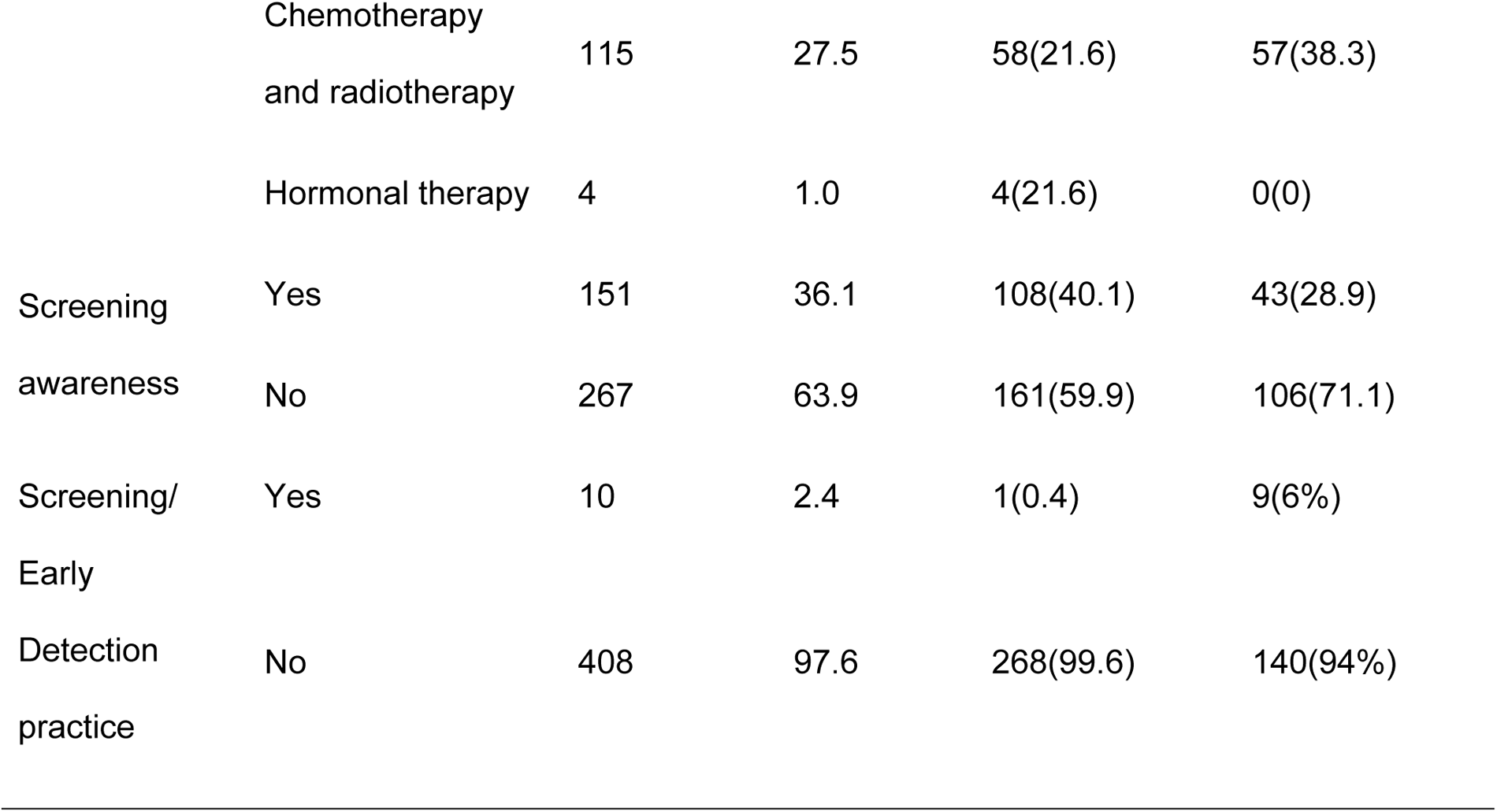
Clinical characteristics of respondents in TASH and SPHMMC, Addis Ababa, Ethiopia 2021.

### Predictors of advanced-stage presentation among breast and cervical cancer patients

Breast and cervical cancer patients who had been referred indirectly were more likely to present at an advanced disease stage than those who had been referred directly (AOR of 3.843; 95%CI = 1.485–9.946). Similarly, rural residents were 2 times more likely to be diagnosed at an advanced disease stages than those who lived in urban areas (AOR of 2.041; 95%CI = 1.108–3.758). Breast and cervical cancer patients who had a patient delay >3 months were 2.5 times more likely to have advanced-stage presentation than their counterparts (AOR = 2.479; 95% CI = 1.078–5.704). Moreover, breast and cervical cancer patients who reported financial difficulties were more likely to have advanced-stage presentation than their counterparts (AOR = 10.102; 95% CI = 1.658– 10.248). A substantial percentage, 73.7%(308) of breast and cervical cancer patients were unaware of the symptoms before being diagnosed, causing the patient to appear late for diagnosis and treatment. Those women who had low awareness about screening and disease itself were approximately 4 times more likely to have advanced-stage presentation than who had higher awareness (AOR = 4.092; 95% CI = 1.078–5.704). Our findings show that 45.3% of participants had a delay in diagnosis following the initial medical consultation. The health system (delayed referral and waiting test results) or the health care provider (misdiagnosis) were responsible for the delay in diagnosis.

**Table 4.** Factors associated with advanced-stage presentation among breast/cervical cancer patients in TASH and SPHMMC Addis Ababa, Ethiopia, 2021.

| Variable | Category | COR with 95% CI | AOR with 95% CI |
| --- | --- | --- | --- |
| Referral status | direct | 1.0 | 1.0 |
|  | indirect | 18.079 (10.652-30.683) | 3.843 (1.485-9.946) |
| Financial difficulties reported | No | 1.0 | 1.0 |
|  | Yes | 4.887 (1.125-21.226) | 10.248 (1.859-56.495) |
| Awareness level | Lower | 1.0 | 1.0 |
|  | Higher | 17.836 (9.569-33.244) | 4.092 (1.658-10.102) |
| Patient delay >3 months | No | 1.0 | 1.0 |
|  | Yes | 2.943 (1.697-5.101) | 2.479 (1.078-5.704) |
| Residence | Urban | 1.0 | 1.0 |
|  | Rural | 2.580 (1.657-4.015) | 2.041 (1.108-3.758) |

*\*Only variables with $P < 0.25$ were included in the multivariable analysis*

## Discussion

This study examined the predictors of advanced-stage presentation among breast and cervical cancer patients in Ethiopia. About two thirds of breast and cervical cancer patients were diagnosed at an advanced disease stage. An indirect referral path, financial difficulty, rural residence, a low awareness level and a patient delay >3 months were independent predictors of advanced-stage presentation.

The findings of such high proportions of advanced breast cancer disease at diagnosis were consistent with those of north-east Ethiopian (65.7%), Nigerian (67.7%) and Indian studies (68%) [12,17,18]. However, they were lower than those reported in a study conducted in north-west Ethiopia (71.2%)[3] and higher than those of studies conducted in South Africa (51%), Pakistan (59%) and Germany (51.6%)[19–21]. The proportion of cervical cancer patients with an advanced stage of disease was nearly similar to those in studies conducted in Nigeria (72.8%)[15] and Sudan (72%)[22] and higher than that in a study conducted in TASH in 2021, which was (56.8%)[23], and a population-based cross-sectional study conducted in Addis Ababa (60.4%)[24].

According to the findings, a large proportion of women with breast and cervical cancer were unaware of the symptoms prior to their diagnosis, resulting in a delay in patient presentation for diagnosis and treatment. The proportion of patients who were delayed accounts for 73.7% (308), which is higher than that in studies conducted at Dessie Referral Hospital, north-east Ethiopia’s only oncology unit 50.5% [25] and northern India 65.3%[12]. Another study from Ethiopia at TASH in 2010 showed that almost half of the study participants (n = 33; 47.8%) said that women in Ethiopia typically know nothing about breast cancer and, in fact, have often never heard of the disease at all. This difference might be due to the difference in sample size of the studies, adoption of different cut-off points for classifying patient delay and a difference in study populations [26].

Our study showed the effects of a patient delay of >3 months on advanced-stage diagnosis of breast and cervical cancer patients. The longer the patient delay time, the more likely the patient was diagnosed at an advanced disease stages. Patients who had a presentation delay of >3 months were 2.4 times more likely to be diagnosed at an advanced stage than those who had no patient delay. Disease progression is expected during a delay. A similar finding was reported in studies conducted in north-east Ethiopia, in which 50.5% (103) women with breast cancer were delayed for >3 months. Out of these delayed women, 75 (72.8%) were diagnosed at a late stage (p < 0.030) [25]. Similarly, a longer patient interval in patients with cervical cancer is associated with more advanced FIGO stages at diagnosis, according to a study performed at TASH [14].

According to our findings, the delay in diagnosis after the initial medical consultation was attributed to either the health system (delayed referral and waiting test result) or the health care provider (misdiagnosis), in 45.3% of cases. A similar result was reported from Egypt (37%) [27].

The study revealed that the odds of advanced-stage presentation were 2.0 times greater in patients residing in rural than in urban areas. This finding is nearly identical to that of a 2021 study in TASH [23] and similar to those of studies conducted in Sudan [22], Nigeria [17] and north-west Ethiopia [3]. This is probably because women who live in places with limited access to health care or who travel longer distances for treatment are more likely experience a delay. Additionally, cancer literacy is probably lower in rural settings. However, this finding contrasts with those of a study on cervical cancer patients conducted in 2019 at TASH Ethiopia [14], which found that the place of residency was not a predictor of advanced-stage disease presentation. This discrepancy could be explained by differences in study methods, settings, sample sizes and research participants.

Our study found no evidence of associations between level of education, age or marital status and advanced-stage presentation. This finding is supported by previous studies in Ethiopia and Nigeria, where age was not associated with an advanced disease stage [14,27]. Nonetheless, this result is in contrast with another studies in Nigeria and Ethiopia, which reported that age and level of education were strong determinants of stage at diagnosis in cervical cancer [23,27]. Women without a high school diploma and those who were above the age of 60 had a considerably higher risk of developing advanced-stage disease than their counterparts [23,27,28]. This disparity could be related to differences in the study location and participants.

Consistent with findings from prior studies [16,19], indirect referral significantly affected advanced-stage presentation in the current study. The odds of advanced-stage presentation were higher among women with breast and cervical cancer patients who were referred indirectly (AOR 3.843, 95% CI (1.485–9.946); this probably reflects the longer duration of referral.

Our study has some limitations. First, even though this study involved a relatively large sample size, due to the nature of a cross-sectional study, the establishment of a causal relationship was not possible. Second, the retrospective nature of information collection from patients on the dates of symptom recognition and medical consultations is prone to recall bias and could have affected the measurement of intervals and other variables, but we have tried to minimize it by using standard tools, carefully designing the questionnaire and training the data collectors.

## Conclusion

Despite the efforts made by the government to improve cancer care and patient outcomes, the majority of breast and cervical cancer patients present at an advanced disease stage. Shortening the referral pathways and designing a direct ‘fast track’ referral for diagnosis and treatment could help to improve the delay in diagnosis and treatment initiation. Implementing diagnostic walk-in centres could be an option to achieve this. Awareness-raising activities should target the general population to promote screening for both cancers and facilitate early detection. Special attention must be given to vulnerable segments of the population, such as rural residents and women with low incomes.

## Data Availability

Data are available from the College of Health Sciences, Addis Ababa University Institutional Ethics Committee (contact via) for researchers who meet the criteria for access to confidential data.

## Abbreviations

AAU: Addis Ababa University
TASH: Tikur Anbessa Specialized Hospital
SPHMMC: St. Paul’s Hospital Millennium Medical College
SPSS: Statistical Package for The Social Sciences
AOR: adjusted odds ratio
CI: confidence interval
SD: standard deviation
HCP: healthcare providers
VIF: variance inflation factor
ETB: Ethiopian birr

## Author Contributions

**Conceptualization:** Birtukan Shewarega, Muluken Gizaw, Nigussie Assefa, Sefonias Getachew, Adamu Addissie, Eva Johanna Kantelhardt, Josephin Trabizsch

**Formal analysis:** Birtukan Shewarega,

**Funding acquisition:** Adamu Addissie, Eva Johanna Kantelhardt

**Investigation:** Birtukan Shewarega

**Methodology**: Birtukan Shewarega, Muluken Gizaw, Nigussie Assefa, Sefonias Getachew, Adamu Addissie, Eva Johanna Kantelhardt, Josephin Trabizsch, Abdu Adem Yesufe, Yonas Dandena, Biruck Gashawubeza

**Project administration:** Adamu Addissie, Eva Johanna Kantelhardt

**Supervision:** Muluken Gizaw, Nigussie Assefa, Abdu Adem Yesufe, Yonas Dandena, Biruck Gashawubeza

**Visualization**: Birtukan Shewarega

**Writing – original draft:** Birtukan Shewarega,

**Writing – review & editing, final approval of the manuscript:** Birtukan Shewarega, Muluken Gizaw, Nigussie Assefa, Sefonias Getachew, Adamu Addissie, Eva Johanna Kantelhardt, Josephin Trabizsch, Abdu Adem Yesufe. Yonas Dandena, Biruck Gashawubeza

## Acknowledgements

The authors would like to thank Tigist Birie and Sosina Kassa for their support during data collection.

## References

1. Sung H, Ferlay J, Siegel RL, Laversanne M, Soerjomataram I, Jemal A, et al. Global cancer statistics 2020: GLOBOCAN estimates of incidence and mortality worldwide for 36 cancers in 185 countries. CA Cancer J Clin. 2021;71(3):209–249

2. 2. WHO, IARC. Global cancer observatory: Cancer today. 2020. Available from URL: https://gco.iarc.fr/today/home

3. Tesfaw A, Tiruneh M, Tamire T, Yosef T. Factors associated with advanced-stage diagnosis of breast cancer in north-west Ethiopia: a cross-sectional study. Ecancermedicalscience. 2021;15.

4. Gebremariam A, Addissie A, Worku A, Assefa M, Pace LE, Kantelhardt EJ, et al. Time intervals experienced between first symptom recognition and pathologic diagnosis of breast cancer in Addis Ababa, Ethiopia: a cross-sectional study. BMJ open. 2019;9(11).

5. Sengayi-Muchengeti M, Joko-Fru WY, Miranda-Filho A, Egue M, Akele-Akpo MT, N’da G, et al. Cervical cancer survival in sub-Saharan Africa by age, stage at diagnosis and Human Development Index: A population-based registry study. Int J Cancer. 2020;147(11):3037–48.

6. Zeleke S, Anley M, Kefale D, Wassihun B. Factors associated with delayed diagnosis of cervical cancer in tikur anbesa specialized hospital, Ethiopia, 2019: Cross-Sectional Study. Cancer Manag Res. 2021;13:579.

7. Duche H, Tsegay AT, Tamirat KS. Identifying risk factors of breast cancer among women attending selected hospitals of Addis Ababa city: Hospital-based unmatched case-control study. Breast Cancer: Targets and Therapy. 2021;13:189.

8. Majeed I, Ammanuallah R, Anwar AW, Rafique HM, Imran FJEMHJ. Diagnostic and treatment delays in breast cancer in association with multiple factors in Pakistan. Eastern Mediterranean Health Journal. 2020;26.

9. Pierz AJ, Randall TC, Castle PE, Adedimeji A, Ingabire C, Kubwimana G, et al. A scoping review: Facilitators and barriers of cervical cancer screening and early diagnosis of breast cancer in Sub-Saharan African health settings. Gynecology oncology report. 2020:100605.

10. Sardi A, Orozco-Urdaneta M, Velez-Mejia C, Perez-Bustos AH, Munoz-Zuluaga C, El-Sharkawy F, et al. Overcoming barriers in the implementation of programs for breast and cervical cancers in Cali, Colombia: a pilot model. J Glob Oncol. 2019;5:1–9.

11. Pearson C, Poirier V, Fitzgerald K, Rubin G, Hamilton WJBo. Cross-sectional study using primary care and cancer registration data to investigate patients with cancer presenting with non-specific symptoms. BMJ open. 2020;10(1).

12. Shreyamsa M, Singh D, Ramakant P, Anand A, Singh KR, Mouli S, et al. Barriers to Timely Diagnosis and Management of Breast Cancer: Observations from a Tertiary Referral Center in Resource Poor Setting. Indian J Surg Oncol. 2020;11(2):287.

13. Dobson CM, Russell AJ, Rubin GPJBhsr. Patient delay in cancer diagnosis: what do we really mean and can we be more specific? BMC health services research. 2014;14(1):1–6.

14. Begoihn M, Mathewos A, Aynalem A, Wondemagegnehu T, Moelle U, Gizaw M, et al. Cervical cancer in Ethiopia–predictors of advanced stage and prolonged time to diagnosis. Infect Agent Cancer. 2019;14(1):36.

15. Awofeso O, Roberts AA, Salako O, Balogun L, Okediji P. Prevalence and pattern of late-stage presentation in women with breast and cervical cancers in Lagos University Teaching Hospital, Nigeria. Nigerian Medical Journal: J Nigeria Medical Association. 2018;59(6):74.

16. Pothas C, Nietz S, Oluwa T, Witness M, Cubasch H, Joffe M. Time to first health care contact, referral pathways and stage at presentation–the experience from four South African breast units. Eur J Cancer. 2020;138:S69.

17. Jedy-Agba E, McCormack V, Olaomi O, Badejo W, Yilkudi M, Yawe T, et al. Determinants of stage at diagnosis of breast cancer in Nigerian women: sociodemographic, breast cancer awareness, health care access and clinical factors. Cancer Causes Control. 2017;28(7):685–97.

18. Dye TD, Bogale S, Hobden C, Tilahun Y, Hechter V, Deressa T, et al. Complex care systems in developing countries: breast cancer patient navigation in Ethiopia. Cancer. 2010;116(3):577–85.

19. Joffe M, Ayeni O, Norris SA, McCormack VA, Ruff P, Das I, et al. Barriers to early presentation of breast cancer among women in Soweto, South Africa. PloS one. 2018;13(2):e0192071.

20. Arndt V, Stürmer T, Stegmaier C, Ziegler H, Dhom G, Brenner HJBjoc. Patient delay and stage of diagnosis among breast cancer patients in Germany–a population based study. JournalCancer . 2002;86(7):1034–40.

21. Gulzar F, Akhtar MS, Sadiq R, Bashir S, Jamil S, Baig SM. Identifying the reasons for delayed presentation of Pakistani breast cancer patients at a tertiary care hospital. Cancer Man Res. 2019;11:1087.

22. Ibrahim A, Rasch V, Pukkala E, Aro ARJIjowsh. Predictors of cervical cancer being at an advanced stage at diagnosis in Sudan. Int J Womens Health. 2011;3:385.

23. Wassie M, Fentie B. Prevalence of late-stage presentation and associated factors of cervical cancer patients in Tikur Anbesa Specialized Hospital, Ethiopia: institutional based cross-sectional study. Infect Agent Cancer. 2021;16 (1):1–6.

24. Dereje N, Gebremariam A, Addissie A, Worku A, Assefa M, Abraha A, et al. Factors associated with advanced stage at diagnosis of cervical cancer in Addis Ababa, Ethiopia: a population-based study. BMJ open. 2020;10(10):e040645.

25. Hassen AM, Hussien FM, Asfaw ZA, Assen HE. Factors Associated with Delay in Breast Cancer Presentation at the Only Oncology Center in North East Ethiopia: A Cross-Sectional Study. J Multidiscip Healthc. 2021;14:681.

26. De Ver Dye T, Bogale S, Hobden C, Tilahun Y, Hechter V, Deressa T, et al. A mixed-method assessment of beliefs and practice around breast cancer in Ethiopia: implications for public health programming and cancer control. Glob Public Health. 2011;6(7):719–31.

27. Abdel-Fattah MM, Anwar MA, Mari E, El-Shazly MK, Zaki AA, Bedwani RN, et al. Patient- and system-related diagnostic delay in breast cancer. Eur J Public Health. 1999;9(1):15–9.

28. McKenzie F, Zietsman A, Galukande M, Anele A, Adisa C, Parham G, et al. Drivers of advanced stage at breast cancer diagnosis in the multicountry A frican breast cancer–disparities in outcomes (ABC-DO) study. Int J Cancer. 2018;142(8):1568–79.

